# Systematic Review of Prevalence of Sleep Problems in India: A Wake- up Call for Promotion of Sleep Health

**DOI:** 10.1101/2023.12.29.23300624

**Authors:** Karuna Datta, Anna Bhutambare, Hruda Nanda Mallick

**Affiliations:** Professor and Head, Department of Sports Medicine, AFMC Pune, Maharashtra, India; JRF 1, DST SATYAM, Human Sleep Research Lab, c/o Dept of Sports Medicine, AFMC Pune, Maharashtra, India; Professor and Faculty Medical Sciences, SGT University, Gurugram, Haryana

## Abstract

An ever-increasing burden of non-communicable diseases, especially in the post pandemic times and an association of sleep problems with them highlighted a felt need to estimate the sleep problem in India. A meta-analysis of the studies conducted on Indian data was planned adhering to PRISMA guidelines. An electronic search of available literature was performed on databases including PubMed, Google Scholar, PsycNet, and Epistemonikos. 100 eligible articles were analysed. To assess the methodological quality 10-points Joanna Briggs Institute (JBI) checklist for prevalence studies was used. The pooled estimates for prevalence of Insomnia found were 25.7%, OSA 37.4%, and RLS 10.6%. An increased prevalence was seen in patients of diabetes, heart disease patients and in otherwise healthy population. Subgroup analysis showed a higher prevalence in patient population and in the otherwise healthy population too,; e.g. Insomnia 32.3% (95% CI: 18.6% to 49.9%, I^2^=99.4%) and 15.1% (95% CI: 8.0% to 26.6%, I^2^=99.1%); OSA 48.1% (95% CI: 36.1% to 60.3%, I^2^=97.4%) and 14.6% (95% CI: 9.2% to 22.5%, I^2^=97.4%) and RLS 13.1% (95% CI: 8.7% to 19.3%, I^2^=91.9%) and 6.6% (95% CI: 2.4% to 16.4%, I^2^=99.1%) respectively. Excessive daytime sleepiness remained prevalent (19.6%) (95 % CI: 8.4% to 39.1%, I^2^=99.8%) in the healthy, which was alarming. A multipronged approach for sleep management, evaluation and research is the need of the hour for managing non communicable disorders and for promoting sleep health in the healthy population.

## 1. Introduction

Understanding the burden of sleep problems on a community is the first step to ensure that the system gears up to take control. Sleep problems and disorders are widely prevalent and continue to emerge especially in the post pandemic world^1^. Establishing prevalence of a problem in the country can help and devise strategies to counter them. A need to develop strategies to combat sleep problems and reduce the burden of these disorders is vital^2^. There was a felt need to analyse the prevalence of sleep problems in India from the available published literature. Moreover, there is an increasing burden of non-communicable diseases. Sleep plays an important role in the pathophysiology of these diseases. Sleep disorders add an allostatic load and hamper the progression of these diseases.

## 2. Material and Methods

’Meta-analysis of epidemiological studies on prevalence of sleep disorders in Indian population’ Trial was registered with Prospero (ID-CRD42022368993).

### 2.1 Search Strategy

The Preferred Reporting Items for Systematic Reviews and Meta Analysis (PRISMA) protocol 2021 guidelines were followed for this systematic review ^3^. An electronic search of available literature was performed on databases including PubMed, Google Scholar, PsycNet, and Epistemonikos. The search keywords consist of various sleep diseases with Boolean Operator (OR) and Boolean Operator (AND) India to combine studies only from India was used. We applied no language restriction and no publication time restriction. Detailed search strategy is provided in **supplementary material (S1)**.

### 2.2 Eligibility Criteria and Study Selection

All the studies conducted on humans in the region of India reporting prevalence of any sleep disorder was included. We included all kinds of studies conducted on various participants (any age group, patients, or general population.). Exposure to any kind of sleep related disorders like-Insomnia, hypersomnia, parasomnia, sleep apnea, sleep paralysis, restless leg Syndrome (RLS), narcolepsy, snoring, chronic fatigue syndrome, seasonal affective disorder, REM sleep behaviour disorder, non-REM sleep behaviour disorder, excessive daytime sleepiness (EDS), periodic limb movement, sleep talking, sleep terror, sleep related breathing disorder, circadian rhythm sleep disorder was considered. Titles and abstracts of the available studies were assessed by two reviewers independently to check if they met the inclusion criteria.

### 2.3 Data Collection

The systematic review was done for the available literature to study the prevalence of sleep disorders in India. The initial literature search resulted in 1802 relevant articles. Of the 1802 articles, 152 articles were initially found eligible. Articles without full text and those in duplicate were removed. PubMed search provided 1200 articles, after screening only 90 articles which reported relevant data were included. PsycNet search produced 202 articles. None of these articles were eligible after screening. Google scholar search provided 250 articles, out of these 37 reported relevant data but 30 articles were repeated thus only 7 articles were included. Epistemonikos search provided 150 articles, only 3 were relevant and finally included. Thus, a total of 100 relevant articles were used for final analysis. The study selection process is shown in **Figure 1**.

**Figure 1:**
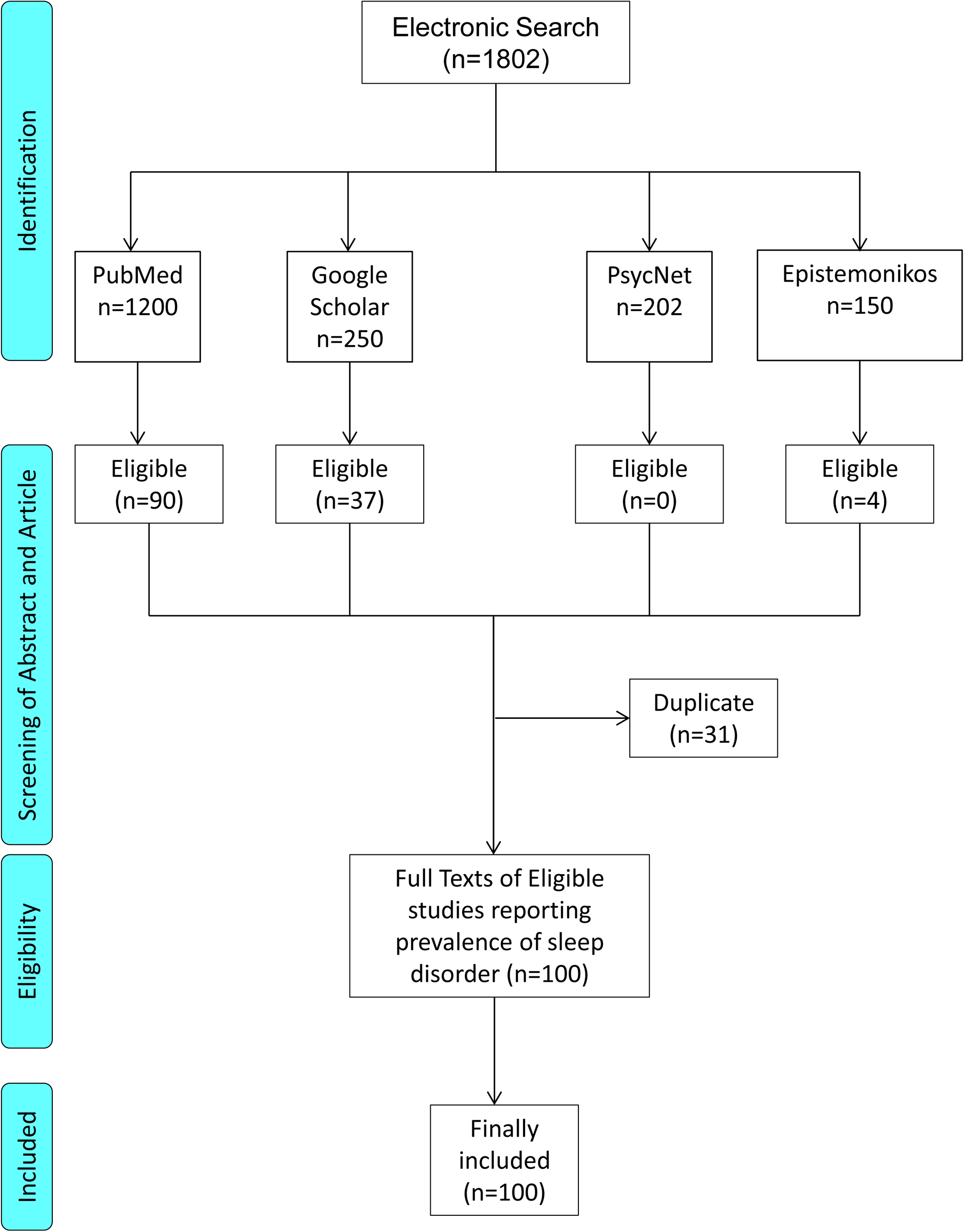
Flow chart of studies screened and included.

### 2.4 Data Extraction

After removing duplicate studies, full text articles were reviewed and following data was extracted - Author name, year of publication, study title, study design, type of study participants, number of participants, age group, disease addressed in the study, number of events or proportion of cases, scale used for disease assessment. If relevant data was not available or full text article was not freely available, then corresponding authors were contacted by email and were requested to provide full text article or relevant data. The relevant data was manually checked by one author and was rechecked by the other author independently. Final table prepared showing detailed characteristics of the included studies is placed in **Table 1**.

### 2.5 Quality assessment of included study

The quality assessment of the included studies for prevalence was done using Joanna Briggs Institute (JBI) checklist ^4^. It consists of ten questions/checklist points with response to each as either ‘yes’, ‘no’, ‘unclear’ or not applicable’. For each ‘yes’, the study is rewarded 1 point otherwise zero and hence a cumulative score 0-10 is allotted. Quality assessment was done independently by two reviewers (AB and KD) and in case of any discrepancy third reviewer was asked. Based on JBI appraisal tool total scores were calculated and articles were assigned quality points.

### 2.6 Data Analysis

A random-effect model was used with inverse variance method for estimating the pooled prevalence or the overall prevalence of some common sleep disorders. We used transformed proportion using logit transformation instead of raw proportion values. The between-study heterogeneity variance was estimated using τ^2^ (tau-squared) and I^2^ statistic with restricted maximum-likelihood estimator method.

Also, the prediction interval for future studies was computed to give the expected range of true prevalence in a new study, using the data. To evaluate between study heterogeneity Cochran’s Q-test was done. Subgroup analysis was also done based on participants’ type (Healthy or Patient) and, again based on age group for studies based on healthy participants.

All statistical analysis was conducted in statistical R-software (Version 4.1.2) using the packages ‘meta’ and ‘metafor’.

## 3. Results

100 studies included in the final analysis showed major sleep disorders like insomnia, obstructive sleep apnea (OSA), restless leg syndrome (RLS) on a sample of 67844 individuals. Meta-analysis for each disorder was done. Excessive daytime sleepiness (EDS) remained a symptom which was found in many studies and analysis for EDS was done. Studies reporting insomnia or insomnia like features^5–29^, OSA^13,22,26,27,29–75^, RLS^10,15,19,25,28,76–90^ and excessive daytime sleepiness^11,17,40,59,60,67,72,80,88,91–104^ were analysed. The details of which are tabulated in Table 1.

The studies in relation to insomnia, OSA, RLS and EDS have been tabulated and provided in **Table 1**.

**Table 1** also shows the quality assessment score. The quality score based on JBI appraisal tool for prevalence studies showed a median score was 7 (range: 4-10) points. There were 4 studies with lowest JBI score of 4 and 7 with the highest JBI score of 10.

**Table 1:**
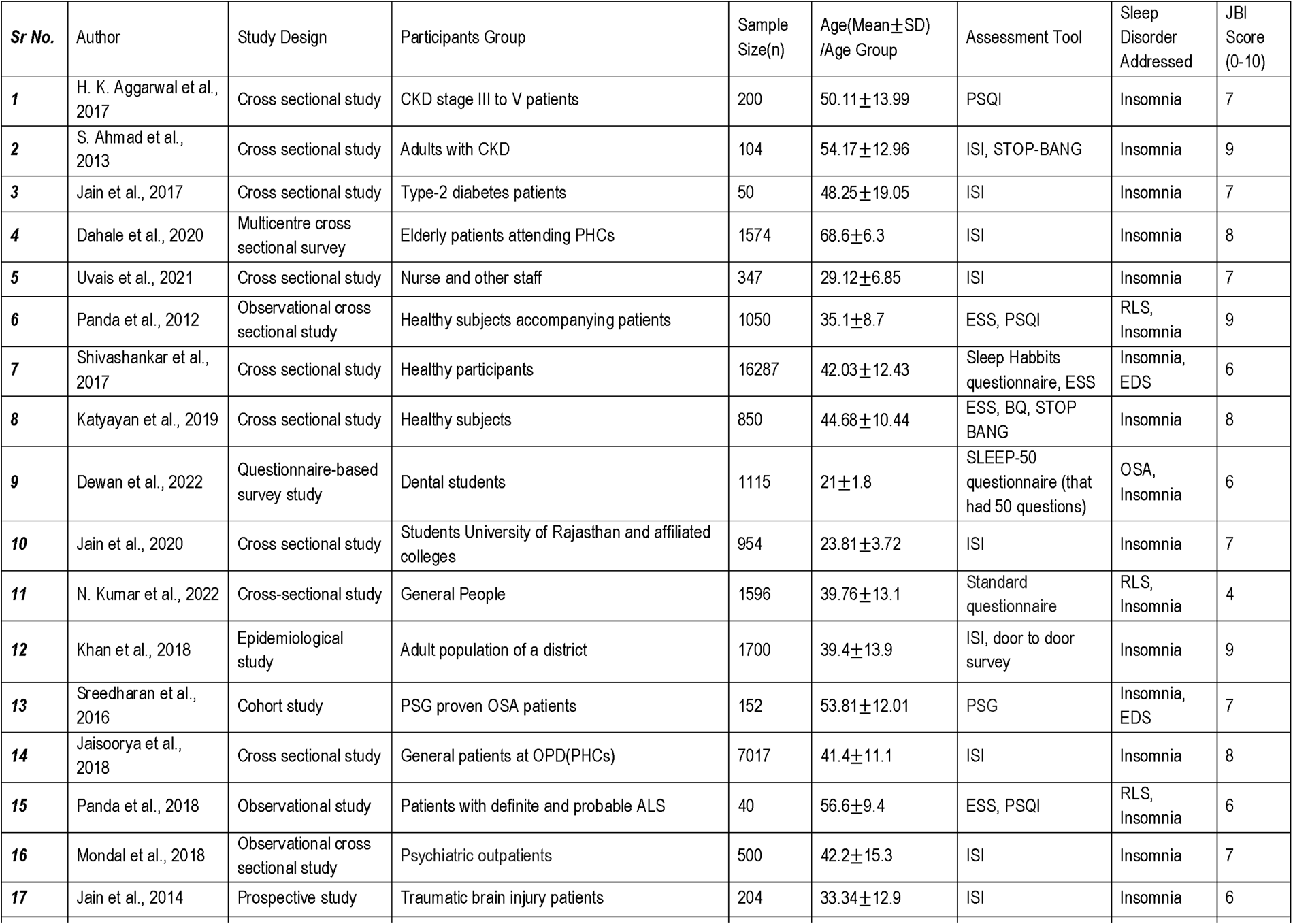
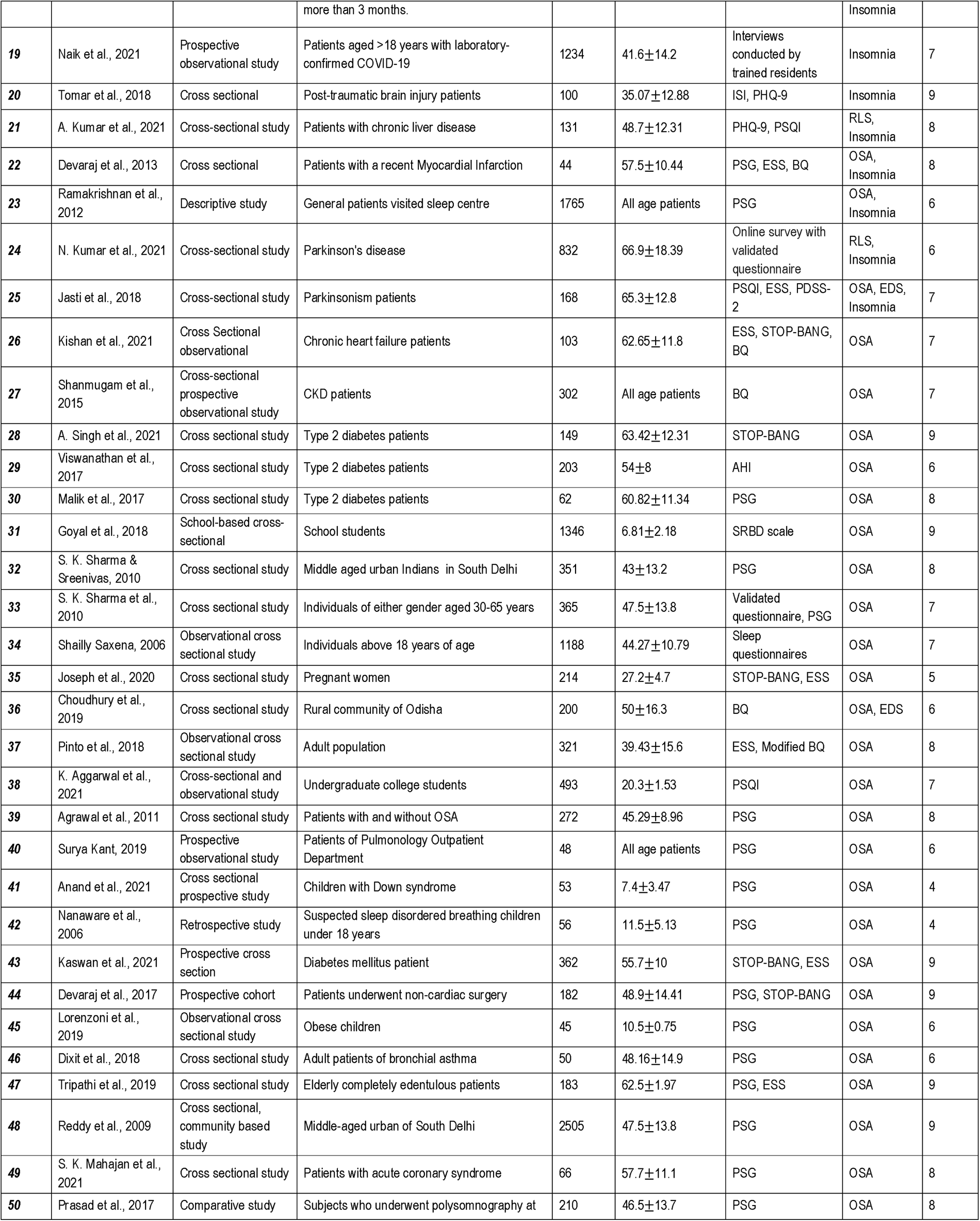
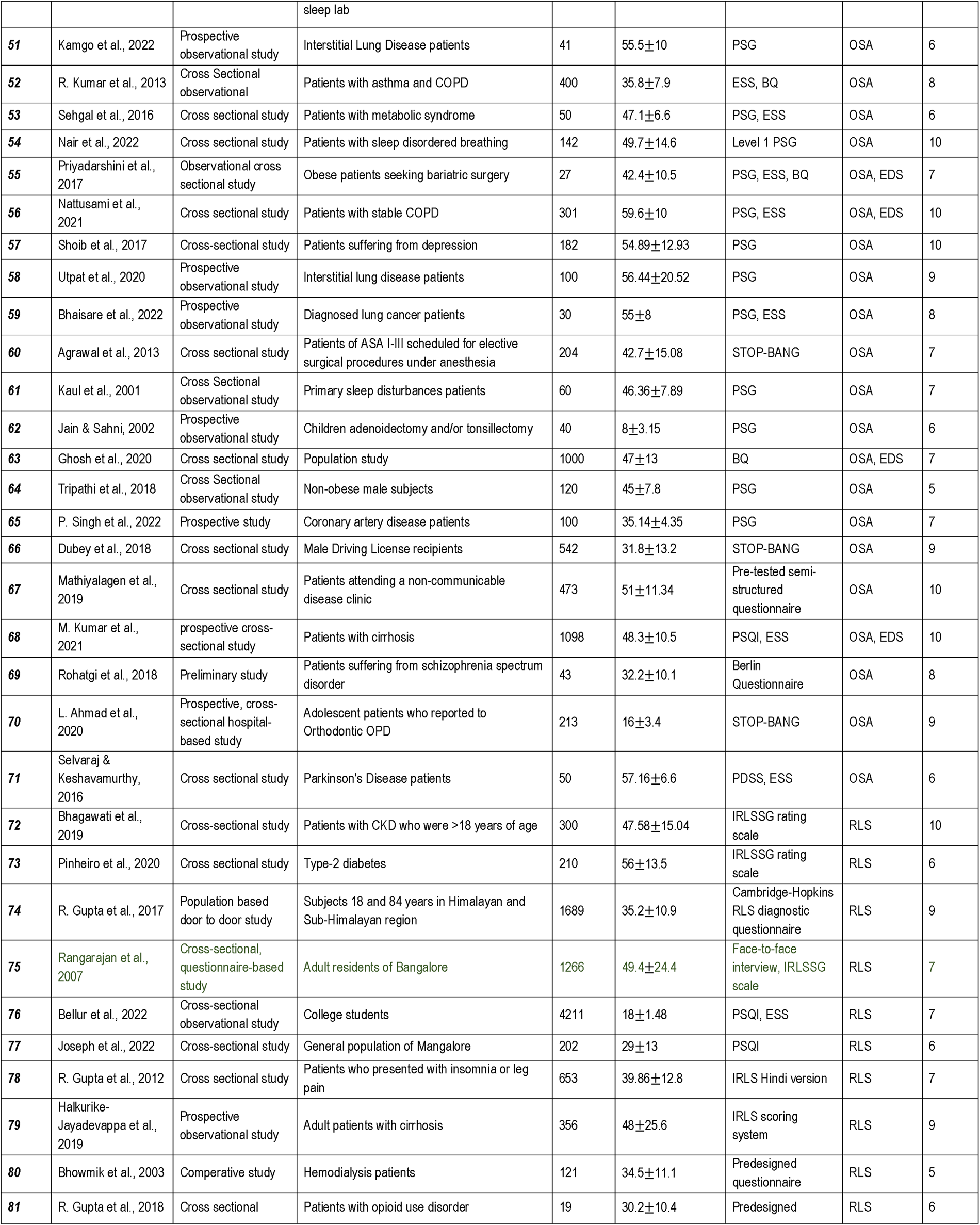
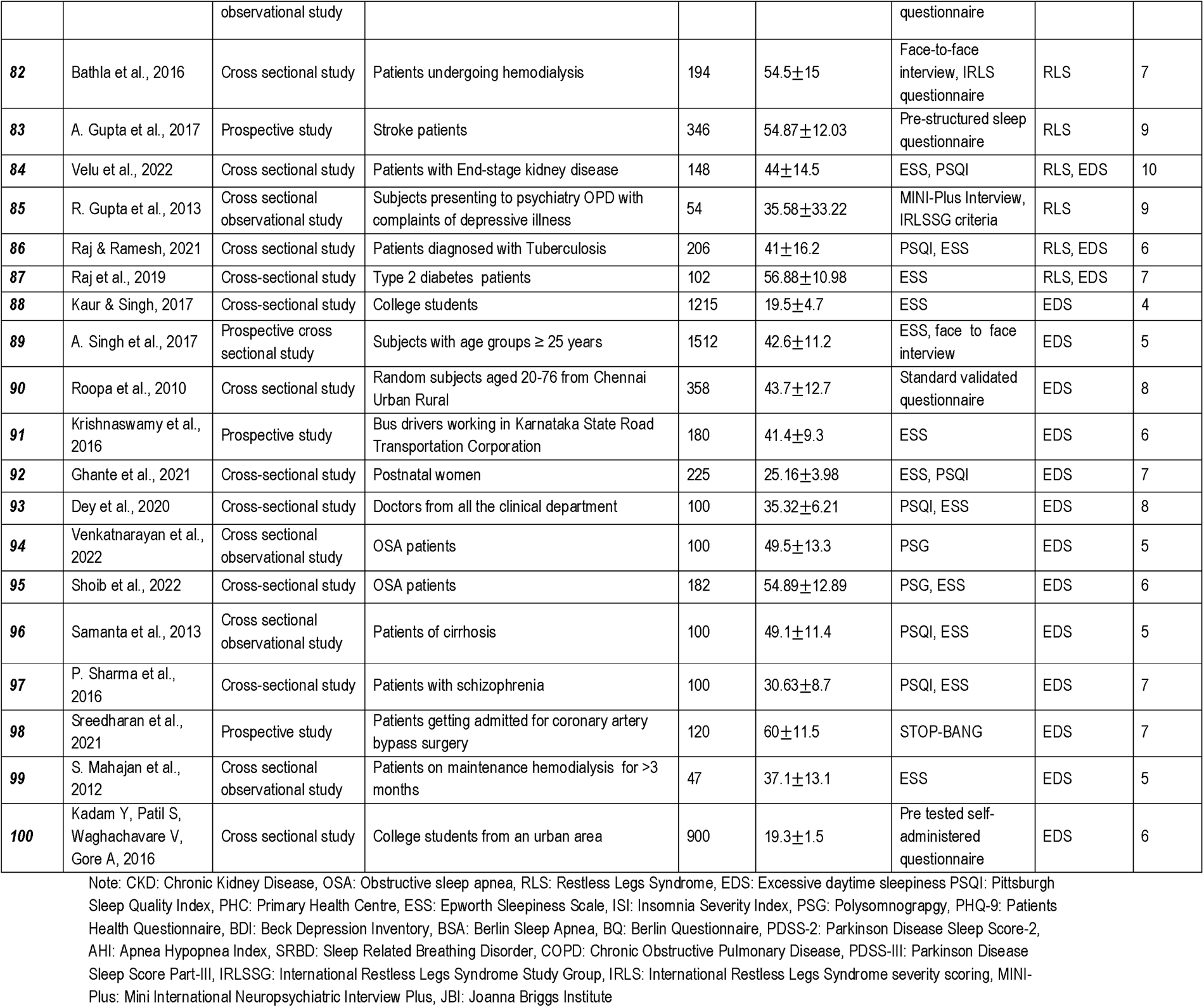
Detailed Characteristics of the Studies Included in the Systematic Review.

A preliminary prevalence of pooled data for insomnia, OSA, and RLS was carried out. The studies showed both healthy and patient population data. Meta-analysis forest plots are shown in **Supplementary Figures 1-4** . Studies were analysed disease wise for general population and patient population and subgroup analysis done for Insomnia, OSA and RLS as shown in **Table 2 (A)**. Subgroup analysis was conducted in healthy population for prevalence of Insomnia, OSA, and RLS as shown in **Table 2 (B)** . Meta-analysis for EDS was also conducted with subgroup analysis based on participant types (healthy and patients) and further based on population group (College students and Adult General Population) in studies with healthy participants as shown in **Table 3**. Forest plots are shown in **Figures 2, 3 and 4** respectively. Some studies showed the prevalence of multiple sleep disorders whereas some only focussed on one of the sleep disorders, so they are included in the disorder or disorders as applicable.

**Table 2:**
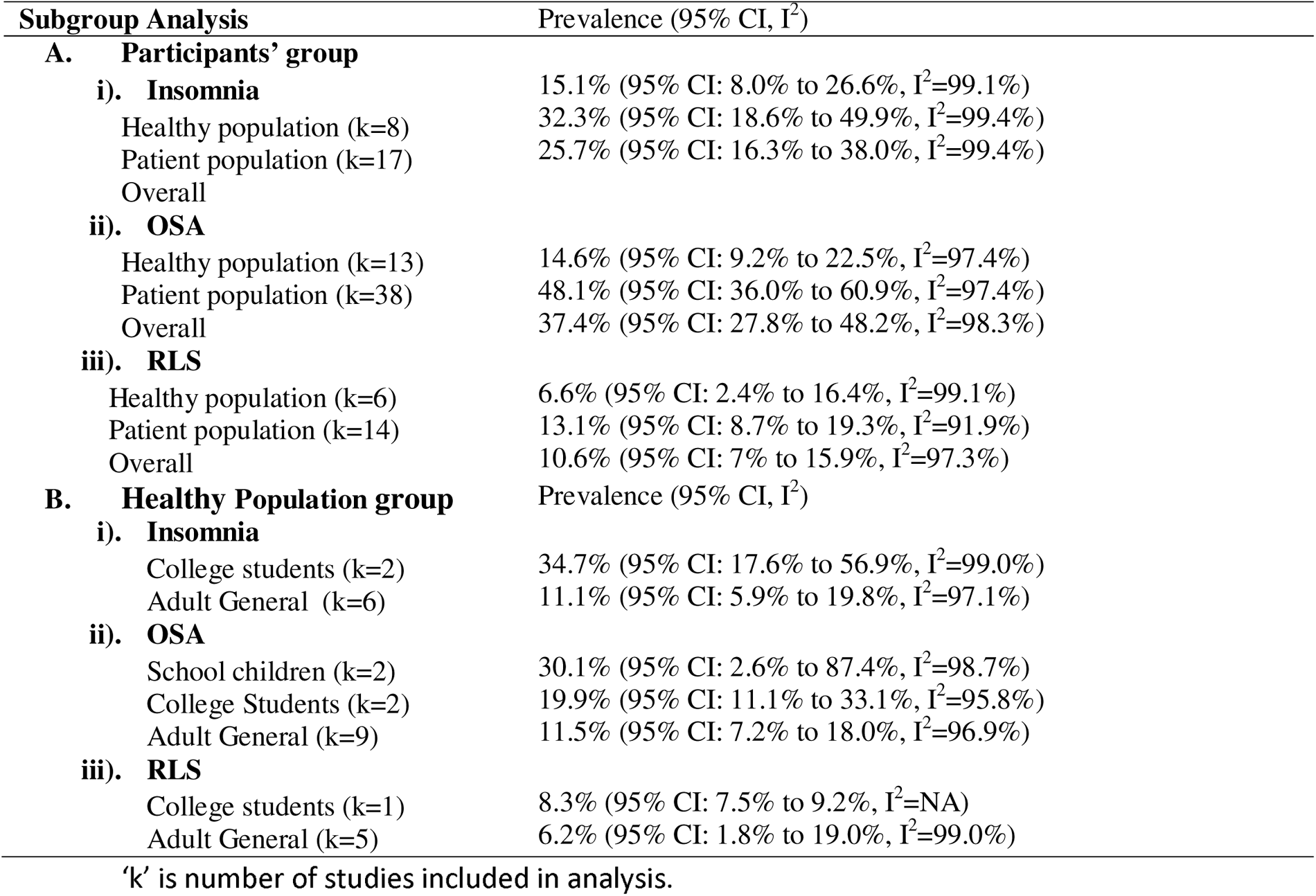
Prevalence of Insomnia, OSA and RLS among Healthy and Patient Population Groups.

**Table 3:** Prevalence of EDS among Patient population and Healthy Population.

| Subgroup Analysis | Prevalence (95% CI, I<sup>2</sup>) |
| --- | --- |
| **A. Participants' Group** |  |
| Healthy population (k=11) | 19.6% (95 % CI: 8.4% to 39.1%, I<sup>2</sup>=99.8%) |
| Patient population (k=14) | 31.5% (95% CI: 20.8% to 44.6%, I<sup>2</sup>=96.5%) |
| Overall | 25.9% (95% CI: 17.1% to 37.1%, I<sup>2</sup>=99.6%) |
| **B. Healthy Population Groups** |  |
| College students (k=3) | 55.1% (95% CI: 30.8% to 77.2%, I<sup>2</sup>= 99.5%) |
| General people (k=8) | 11.6% (95% CI: 4.6% to 26.2%, I<sup>2</sup>=99.4%) |

'k' is number of studies included in analysis.

**Figure 2:**
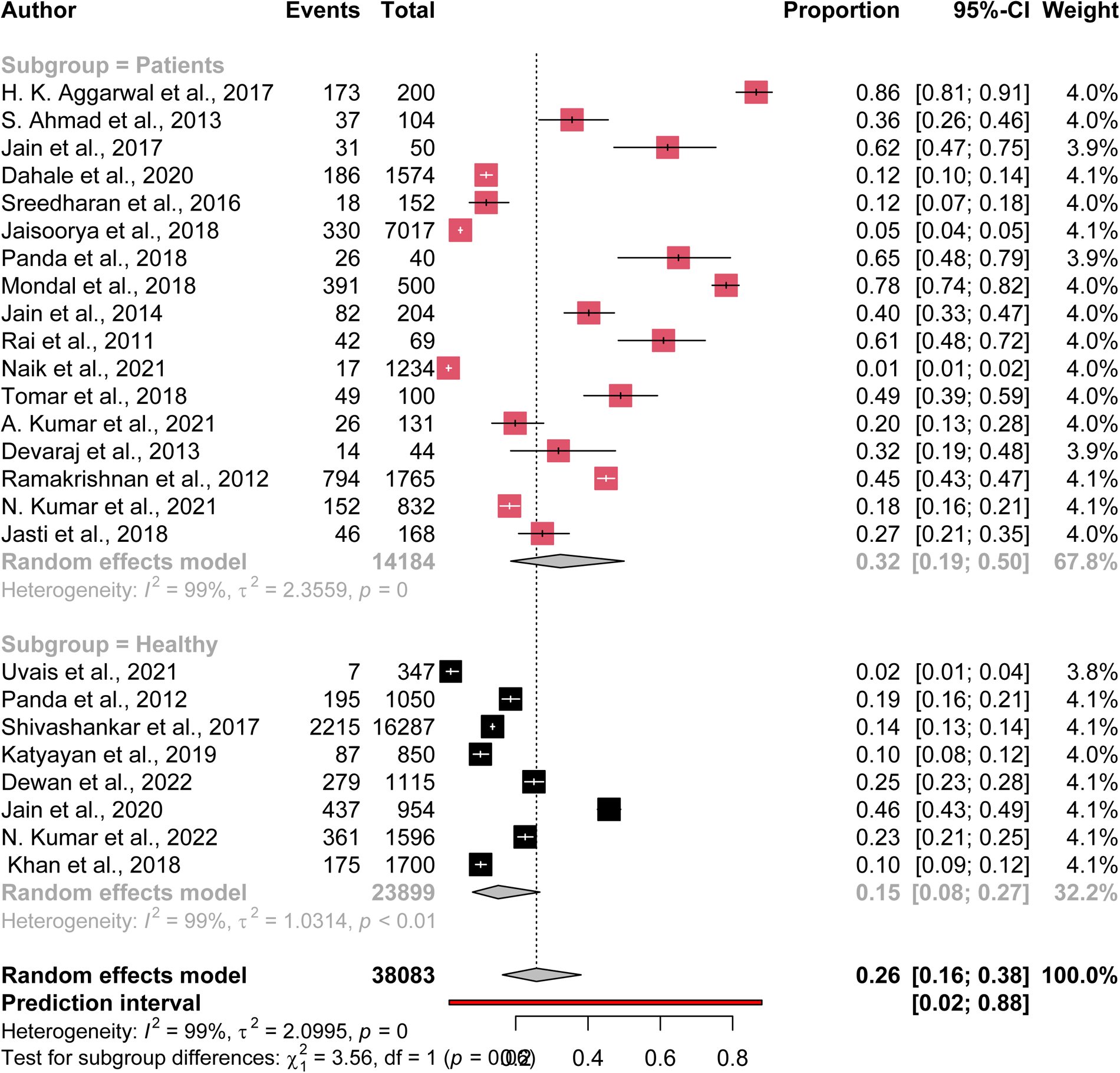
Forest plot of subgroup analysis-Insomnia.

**Figure 3:**
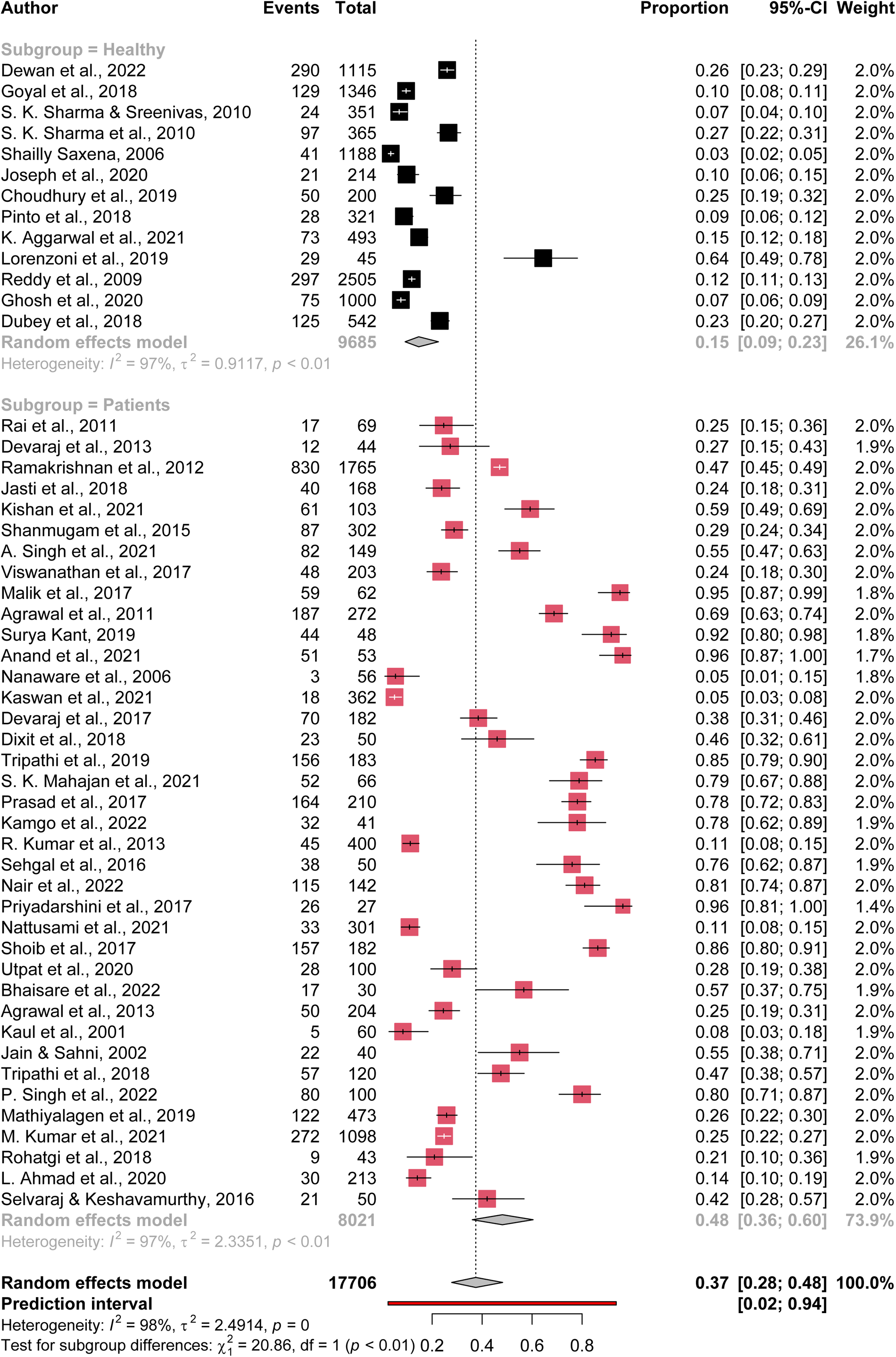
Forest plot of subgroup analysis-Obstructive Sleep Apnea.

**Figure 4:**
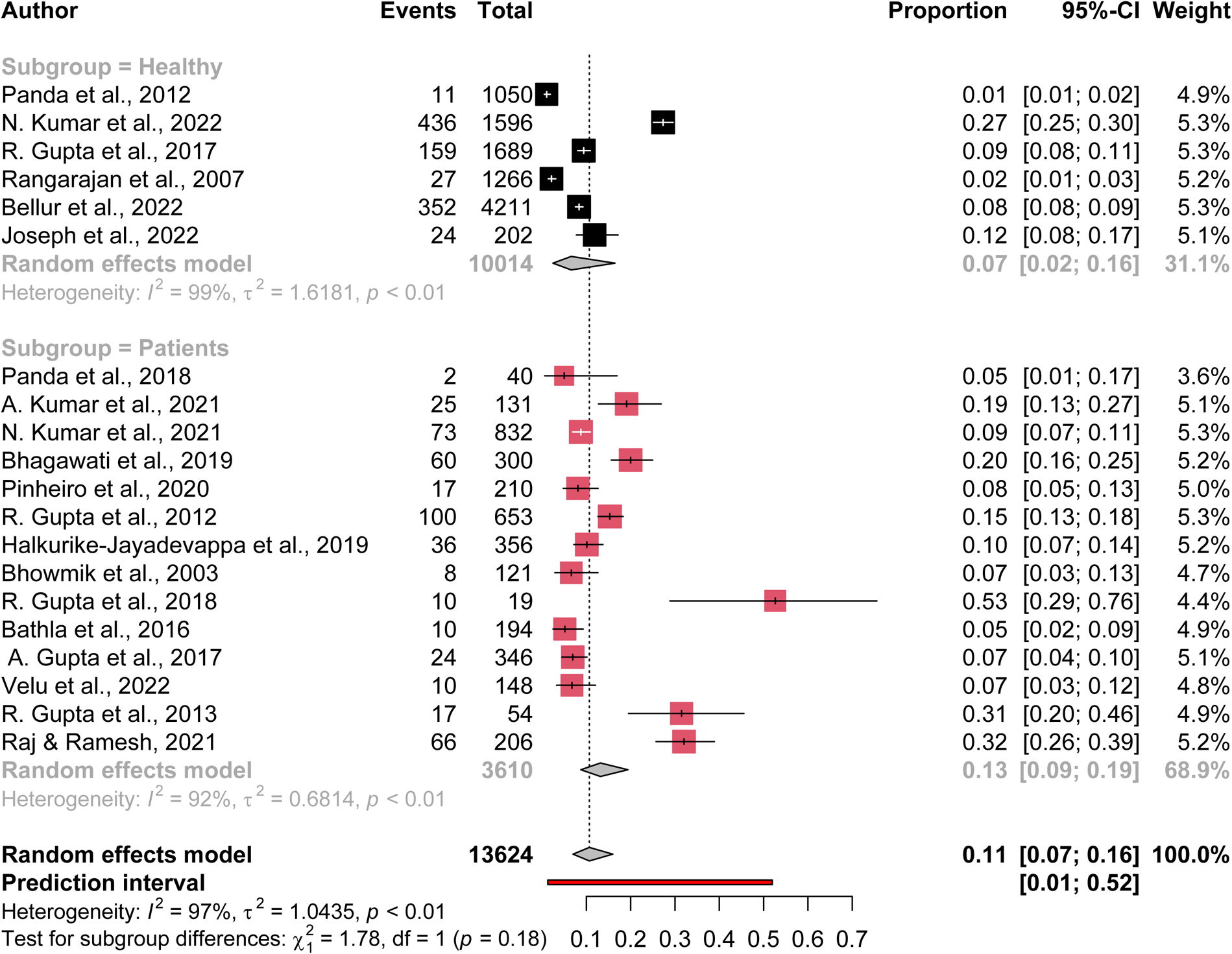
Forest plot of subgroup analysis-Restless Leg Syndrome.

## 4. Discussion

The studies analysed showed that the patient data was being collected from outpatient department, so the patient had a morbidity for which they had reported. Some were done on caregivers and attendants^9,10,97^ of patients while they were in hospital or attending on a patient while in outpatient department. Few studies worked on general population using representative samples of college students^13,14,35,42,80,92,104^ and some targeted the population in general ^12,16,37,38,40,67,78,79,81,93,94^.

A pooled data of both healthy and patient population together hints that the prevalence of sleep disorders is very high namely for insomnia being 25.7% (95% CI: 16.3% to 38.0%, I^2^=99.4%), OSA 37.4% (95% CI: 27.8% to 48.2%, I^2^=98.3%) and for RLS 10.6% (95% CI: 7% to 15.9%, I^2^=97.3%). It was found that the methodology implemented in the study also varied widely from surveys, face to face interviews and only few with subjective and objective parameters both. Keeping the large variance in the target population studied and the methodology adopted, the range of prevalence of sleep disorders thus had an increased range. Further subgroup analysis of general population was done to understand the prevalence in an otherwise healthy population.

We had done an analysis of prevalence of insomnia reported by various articles and our results showed an overall of 25.7 % with a high heterogeneity (I^2^=99.4%). The high heterogeneity might have arisen both because of the methods used and the target population studied. The relative high prevalence of insomnia in patients of diabetes is glaring. This requires insomnia treatment guidelines incorporated in the comprehensive approach of dealing with such disorders. Considering the economic burden of non-communicable diseases like diabetes and hypertension in India, a holistic approach to better sleep in such disorders is warranted^105,106^. To ensure that the primary care physician can help in early diagnosis, management, and treatment of insomnia, it is imperative to teach basics of sleep management in MBBS curriculum. A glaring need exists for the primary care physician to be acquainted with the standard treatment guidelines^107^.

Further analysis on healthy participants for insomnia showed a prevalence of 34.7% (95% CI: 17.6% to 56.9%, I^2^=99.0%) in college students and 11.1% (95% CI: 5.9% to 19.8%, I^2^=97.1%) in general population. A relatively high prevalence in college students raises an alarm. This needs to be reformed to ensure a healthier future population. College going students should learn ill effects of sleep deprivation and importance of addressing their sleep issues earlier to avoid the resurgence of somatic complaints and psychiatric symptoms in later life. Education and self-motivation remain the cornerstone of this strategy. In fact, this education should start at a very young age. A systematic review on the effects of the pandemic on sleep in children and adolescents highlights the growing problem ^108,109^ . There is a need to develop sleep hygiene principles, like dental hygiene, right from preschool and nursery days. Study on urban school children mentioned the role parents play in setting bedtimes. The study brought out that the high school children felt that parental control was much less and that it diminished from middle school onwards ^110^. Parental awareness to improve control over sleep times during both weekdays and weekends is vital. OSA has been studied in various diseases like diabetes, chronic heart failure, Parkinson’s disease, psychiatric disorders, cirrhosis, etc. Data also exists about its prevalence in general population. Methods employed ranged from questionnaires^5,6,8–10,12–14,16,18–21,24,25,29–32,34,38–40,42,44,47,48,51,52,55,56,59,60,63,64,67,70–75,80,82,84,87,88,90–97,99–104^, home testing to overnight sleep studies^34,49,51,52,55,58–66,68,69,98,99^. In some studies, screening using questionnaires was done followed by polysomnography^10,26,48,51,52,59,64,99^. Our analysis of 51studies available on India data showed a prevalence of 37.4% (95% CI: 27.8% to 48.2%, I^2^=98.3%). There were a high heterogeneity of the study results due to different age groups, health status level i.e., healthy, and patient groups, different study designs, methodology and even outcome parameters. The prevalence varied from 9.2% to 22.5% in general population. A similar result of prevalence of 11% was reported in healthy Indian population by TM Suri et al^111^ in a systematic review conducted by them. A relatively higher prevalence of 48.1% was seen in specific patient groups in our study. Considering a sample of 17706 of a heterogeneous group, the least prevalence was in a study done using baseline questionnaires on a healthy population which was also 3%! In a questionnaire based study in Morocco, 13.9% had witnessed apnea ^112^. OSA prevalence studies worldwide showed a wide variability in the type of outcome and instrument used. Studies also utilised screening questionnaires followed by sleep studies^52,113,114^. Questionnaire based studies in Thailand reported 58.5% snoring and 4.8% prevalence of snoring and OSA respectively^114^. In some studies, an increased association with BMI and Adjusted neck circumference was found.

Needless to say, that the disease burden exists at all age groups and hence a comprehensive strategy will benefit not only in overcoming the specific problem but will also help avert the onset of co morbidities like cardiovascular disorder, stroke, metabolic syndrome, etc. which are often associated with OSA. A study done in 2016 showed that the older population who were having witnessed apnea had more severe OSA^102^. Here is the need to prevent the build-up of the disease for more productive and disease-free life later. Strategies aiming on prevention, regular screening, and treatment of the affected population even in its milder forms might reduce this burden. An analysis of prevalence of snoring itself is alarming by its presence in 6–12-year-olds, medical undergraduates and in population-based studies. At the first place why one should be snoring as a child or young adult? Possible reason might be a relative lack of awareness amongst the parents and young adults. Also at the primary care level, even if the patient does complain about snoring, there is lack of availability of what to do in the form of guidelines at the National level. Hence awareness campaigns at schools and colleges and empowering our primary care with standard guidelines is the need of the day.

After the COVID pandemic, sleep management probably does not remain the domain of a sleep expert alone but contribution from every healthcare professional is required. Enhancing sleep education in healthcare curriculum and ensuring that the referral systems are developed for professional handling of the disorder is required.

Excessive daytime sleepiness (EDS) also remained an important symptom to be analysed because of the repercussions of the symptom of safety issues at work, health and economy of the society and hence was also taken separately for subgroup analysis. EDS was found to be associated with OSA, in insomniacs and in RLS patients. Various instruments like surveys, face to face interviews were used to study prevalence. RLS with insomnia was also reported by Kumar et al in Parkinson’s disease patients^28^and RLS with OSA was reported in pulmonary tuberculosis^90^. Using questionnaires, Amyotrophic Lateral Sclerosis patients were found to have insomnia in 65%, snoring in 45% and 5% had a risk of RLS^19^. This makes it an important matter of concern because of the impact of excessive daytime sleepiness on the productivity of the individual, and on safety and health. The presenteeism of an employee can have disastrous consequences. Secondly another point which drives home is the very instruments which help in the calculation of excessive daytime sleepiness; how is it labelled excessive, what are the factors associated with EDS and Is there any role in having a task force to dwell on this concept further since it has repercussions on both health and economy of the society? At a national level, there is a felt need to develop frameworks to promote sleep health by promotive, preventive, curative, and rehabilitative mechanisms. National societies have been doing education programs in sleep for medical colleges, doctors, and technicians. There have been efforts also to raise awareness amongst school children by holding symposiums for school children and for school counsellors. Overall health enhancement using comprehensive public health programs aiming at good sleep promotion may be required.

Though our study has been an extensive review of all the studies relevant to the objective but there are certain limitations. Exact prevalence in the population is still not clear due to a high heterogeneity of the data. Hence, a task force to develop prevalence studies may be deployed for insomnia and OSA. Awareness material developed by sleep experts may be put up on national portals for free access to population.

To conclude, India has a health burden of sleep disorders, and a need to develop strategies to manage them early is imminent. The need to develop standardised protocols to study prevalence of various sleep disorders on a national level remains pivotal.

## Supporting information

Detailed search strategy

Forest Plots of pooled insomnia data

Forest Plots of pooled obstructive sleep apnea data

Forest Plots of pooled Restless Leg Syndrome data

Forest Plots of pooled data of Excessive Daytime Sleepiness

## Acknowledgements

The project was not funded but the meta-analysis was helped by JRF -1 who was appointed under DST SATYAM project scheme.

## Data Availability

The data is available in the manuscript and supplementary files.

