## Supplementary material for "Systematic Review of Prevalence of Sleep Problems in India: A Wake- up Call for Promotion of Sleep Health": Detailed search strategy

Following database were searched for literature available with no language restriction and no publication date restriction.

**1). PubMed Search Result:** Last search on Aug 28, 2022

Search keyword:

((Insomnia) OR (hypersomnia) OR (parasomnia) OR (sleep apnoea) OR (sleep paralysis) OR (restless leg movement) OR (narcolepsy) OR (Snoring) OR (Chronic Fatigue syndrome) OR (seasonal affective disorder) OR (REM Sleep Behaviour Disorder) OR (Non-REM Sleep Behaviour Disorder) OR (Excessive sleepiness) OR (Periodic Limb Movement) OR (Sleep Talking) OR (Sleep Terror) OR (Sleep Related Breathing Disorder) OR (Circadian Rhythm Sleep Disorder) AND (INDIA))

Total results: 1200

Eligible: 194+69=264 Eligible articles

Used for Data Extraction: 132 Articles

**2). PsycNet Search Results:** Last search on Sep 23, 2022

Search Keyword:

Any Field: India AND (Any Field: Insomnia) OR (Any Field: hypersomnia) OR (Any Field: parasomnia) OR (Any Field: sleep apnoea) OR (Any Field: sleep paralysis) OR (Any Field: restless leg movement) OR (Any Field: narcolepsy) OR (Any Field: Snoring) OR (Any Field: Chronic Fatigue syndrome) OR (Any Field: seasonal affective disorder) OR (Any Field: REM Sleep Behaviour Disorder) OR (Any Field: Non-REM Sleep Behaviour Disorder) OR (Any Field: Excessive sleepiness) OR (Any Field: Periodic Limb Movement) OR (Any Field: Sleep Talking) OR (Any Field: Sleep Terror) OR (Any Field: Sleep Related Breathing Disorder) OR (Any Field: Circadian Rhythm Sleep Disorder))

Results: 202 articles

Eligible: None

**3).Google Scholar Search Results:** Last Search on Dec 2022

Search Keyword:

India AND Insomnia OR hypersomnia OR parasomnia OR sleep OR apnoea OR sleep OR paralysis OR restless OR movement OR narcolepsy OR snoring OR bruxism OR REM OR non OR REM OR sleep OR deprivation OR circadian sleep

Results: 250 total

Eligible: 90 (30 repeated).

For Data Extraction: 17 included

**4). Epistemonikos Search Results:** Last search on Dec 2022

Search Keyword:

(title:(India) OR abstract:(India)) AND (title:(Insomnia) OR abstract:(Insomnia)) OR (title:(hypersomnia) OR abstract:(hypersomnia)) OR (title:(parasomnia) OR abstract:(parasomnia)) OR (title:(sleep apnoea) OR abstract:(sleep apnoea)) OR (title:(sleep paralysis) OR abstract:(sleep paralysis)) OR (title:(restless leg movement) OR abstract:(restless leg movement)) OR (title:(narcolepsy) OR abstract:(narcolepsy)) OR (title:(Snoring) OR abstract:(Snoring)) OR (title:(Chronic Fatigue syndrome) OR abstract:(Chronic Fatigue syndrome)) OR (title:(seasonal affective disorder) OR abstract:(seasonal affective disorder)) OR (title:(REM Sleep Behavior Disorder) OR abstract:(REM Sleep Behavior Disorder)) OR (title:(Non-REM Sleep Behaviour Disorder) OR abstract:(Non-REM Sleep Behaviour Disorder)) OR (title:(Excessive sleepiness) OR abstract:(Excessive sleepiness)) OR (title:(Periodic Limb Movement) OR abstract:(Periodic Limb Movement)) OR (title:(Sleep Talking) OR abstract:(Sleep Talking)) OR (title:(Sleep Terror) OR abstract:(Sleep Terror)) OR (title:(Sleep Related Breathing Disorder) OR abstract:(Sleep Related Breathing Disorder)) OR (title:(Circadian Rhythm Sleep Disorder) OR abstract:(Circadian Rhythm Sleep Disorder))

Total Results: 150

Eligible: 4 (1 repeated)

Data extraction: 3 Articles
