## Supplementary figures and images for "Systematic Review of Prevalence of Sleep Problems in India: A Wake- up Call for Promotion of Sleep Health"

### Forest Plots of pooled data of Excessive Daytime Sleepiness

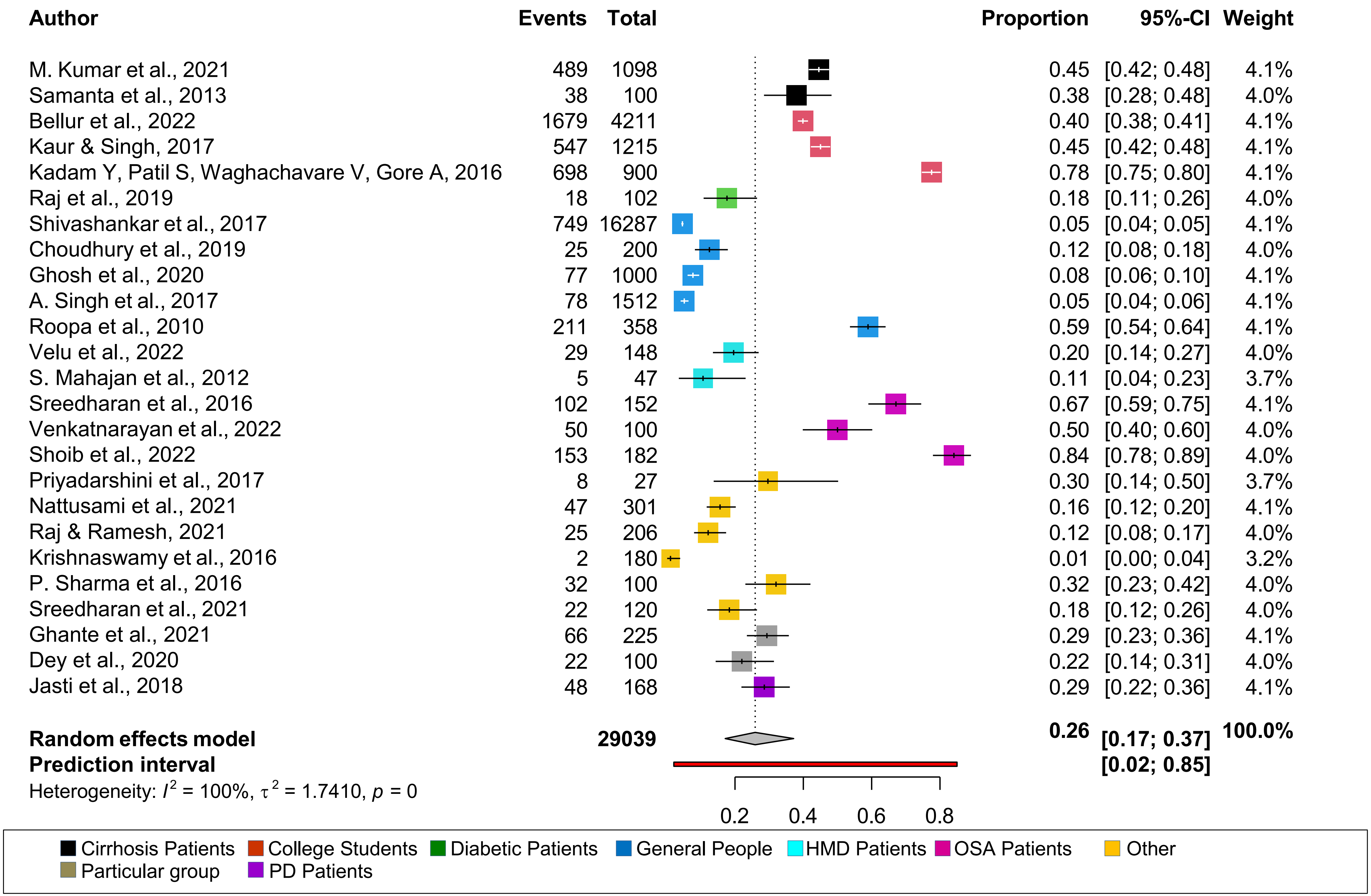

### Forest Plots of pooled insomnia data

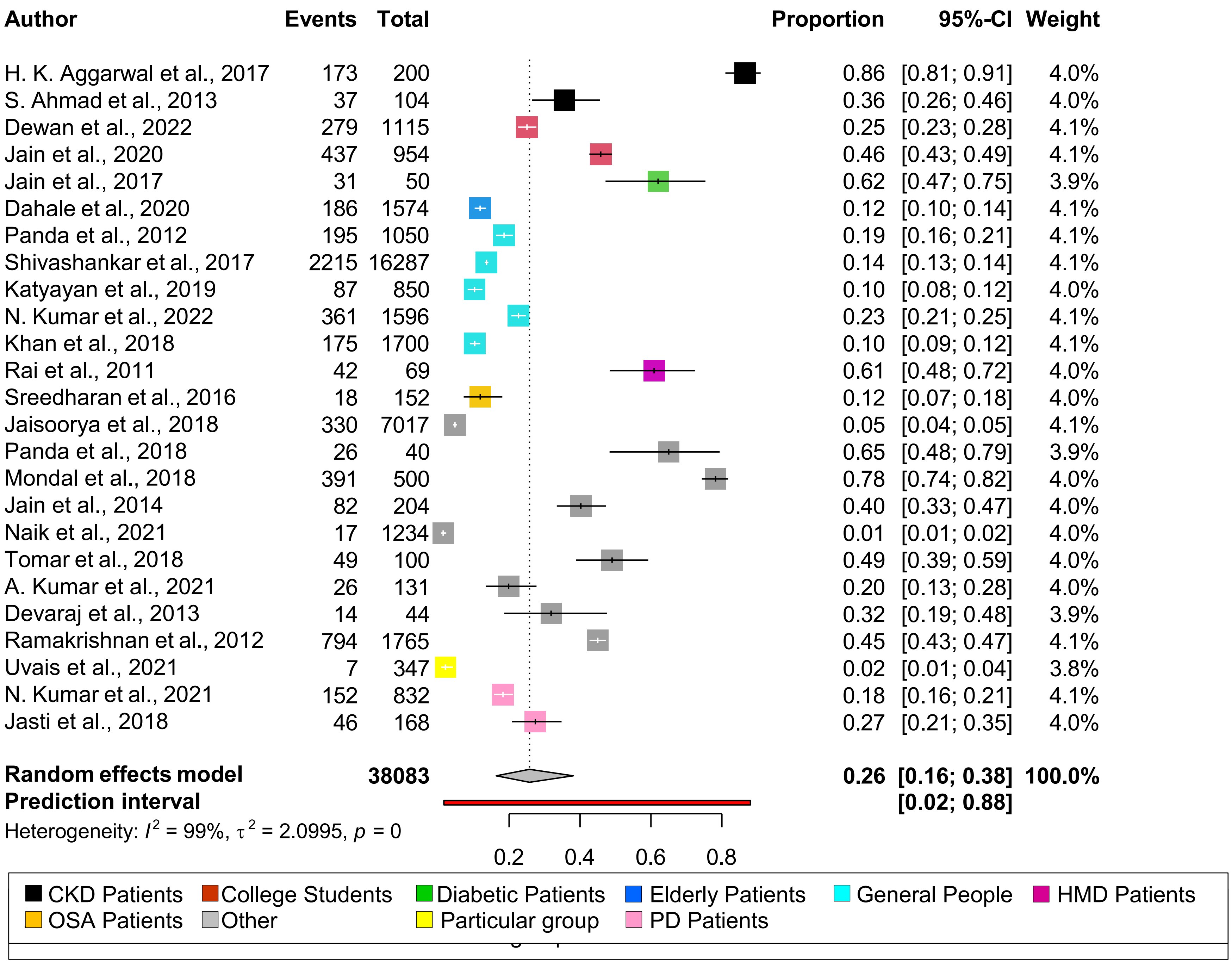

### Forest Plots of pooled Restless Leg Syndrome data

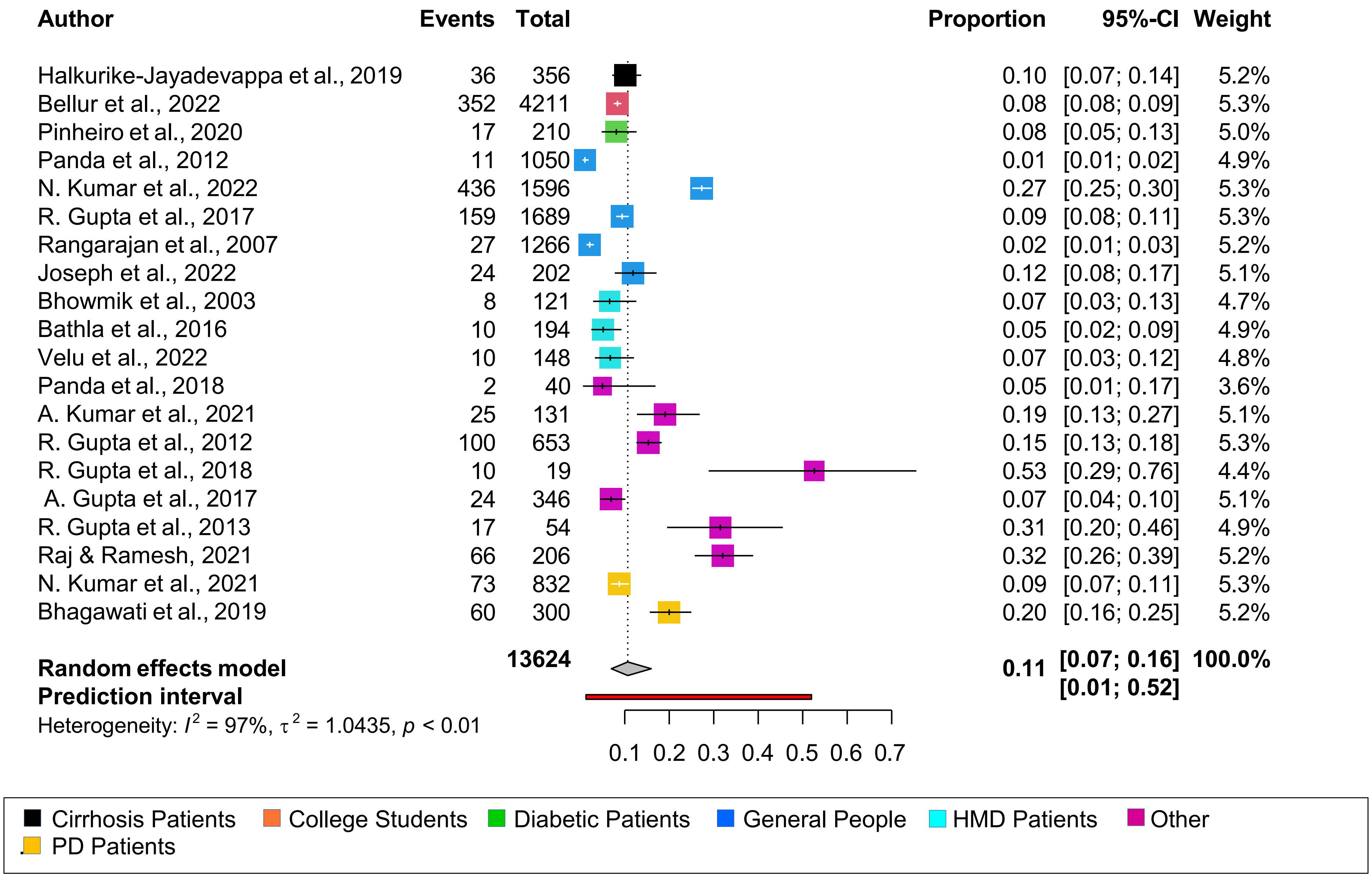
