## Supplementary material for "Systematic Review of Prevalence of Sleep Problems in India: A Wake- up Call for Promotion of Sleep Health": Forest Plots of pooled obstructive sleep apnea data

| Author | Events | Total | Proportion | 95%-CI | Weight |
|---|---|---|---|---|---|
| M. Kumar et al., 2021 | 272 | 1098 | 0.25 | [0.22; 0.27] | 2.0% |
| Shanmugam et al., 2015 | 87 | 302 | 0.29 | [0.24; 0.34] | 2.0% |
| Dewan et al., 2022 | 290 | 1115 | 0.26 | [0.23; 0.29] | 2.0% |
| K. Aggarwal et al., 2021 | 73 | 493 | 0.15 | [0.12; 0.18] | 2.0% |
| A. Singh et al., 2021 | 82 | 149 | 0.55 | [0.47; 0.63] | 2.0% |
| Viswanathan et al., 2017 | 48 | 203 | 0.24 | [0.18; 0.30] | 2.0% |
| Malik et al., 2017 | 59 | 62 | 0.95 | [0.87; 0.99] | 1.8% |
| Kaswan et al., 2021 | 18 | 362 | 0.05 | [0.03; 0.08] | 2.0% |
| S. K. Sharma & Sreenivas, 2010 | 24 | 351 | 0.07 | [0.04; 0.10] | 2.0% |
| S. K. Sharma et al., 2010 | 97 | 365 | 0.27 | [0.22; 0.31] | 2.0% |
| Tripathi et al., 2019 | 156 | 183 | 0.85 | [0.79; 0.90] | 2.0% |
| Shailly Saxena, 2006 | 41 | 1188 | 0.03 | [0.02; 0.05] | 2.0% |
| Choudhury et al., 2019 | 50 | 200 | 0.25 | [0.19; 0.32] | 2.0% |
| Pinto et al., 2018 | 28 | 321 | 0.09 | [0.06; 0.12] | 2.0% |
| Reddy et al., 2009 | 297 | 2505 | 0.12 | [0.11; 0.13] | 2.0% |
| Ghosh et al., 2020 | 75 | 1000 | 0.07 | [0.06; 0.09] | 2.0% |
| Rai et al., 2011 | 17 | 69 | 0.25 | [0.15; 0.36] | 2.0% |
| Agrawal et al., 2011 | 187 | 272 | 0.69 | [0.63; 0.74] | 2.0% |
| Devaraj et al., 2013 | 12 | 44 | 0.27 | [0.15; 0.43] | 1.9% |
| Ramakrishnan et al., 2012 | 830 | 1765 | 0.47 | [0.45; 0.49] | 2.0% |
| Kishan et al., 2021 | 61 | 103 | 0.59 | [0.49; 0.69] | 2.0% |
| Surya Kant, 2019 | 44 | 48 | 0.92 | [0.80; 0.98] | 1.8% |
| Anand et al., 2021 | 51 | 53 | 0.96 | [0.87; 1.00] | 1.7% |
| Nanaware et al., 2006 | 3 | 56 | 0.05 | [0.01; 0.15] | 1.8% |
| Devaraj et al., 2017 | 70 | 182 | 0.38 | [0.31; 0.46] | 2.0% |
| Dixit et al., 2018 | 23 | 50 | 0.46 | [0.32; 0.61] | 2.0% |
| S. K. Mahajan et al., 2021 | 52 | 66 | 0.79 | [0.67; 0.88] | 2.0% |
| Prasad et al., 2017 | 164 | 210 | 0.78 | [0.72; 0.83] | 2.0% |
| Kamgo et al., 2022 | 32 | 41 | 0.78 | [0.62; 0.89] | 1.9% |
| R. Kumar et al., 2013 | 45 | 400 | 0.11 | [0.08; 0.15] | 2.0% |
| Sehgal et al., 2016 | 38 | 50 | 0.76 | [0.62; 0.87] | 1.9% |
| Nair et al., 2022 | 115 | 142 | 0.81 | [0.74; 0.87] | 2.0% |
| Priyadarshini et al., 2017 | 26 | 27 | 0.96 | [0.81; 1.00] | 1.4% |
| Nattusami et al., 2021 | 33 | 301 | 0.11 | [0.08; 0.15] | 2.0% |
| Shoib et al., 2017 | 157 | 182 | 0.86 | [0.80; 0.91] | 2.0% |
| Utpat et al., 2020 | 28 | 100 | 0.28 | [0.19; 0.38] | 2.0% |
| Bhaisare et al., 2022 | 17 | 30 | 0.57 | [0.37; 0.75] | 1.9% |
| Agrawal et al., 2013 | 50 | 204 | 0.25 | [0.19; 0.31] | 2.0% |
| Kaul et al., 2001 | 5 | 60 | 0.08 | [0.03; 0.18] | 1.9% |
| Jain & Sahni, 2002 | 22 | 40 | 0.55 | [0.38; 0.71] | 2.0% |
| Tripathi et al., 2018 | 57 | 120 | 0.47 | [0.38; 0.57] | 2.0% |
| P. Singh et al., 2022 | 80 | 100 | 0.80 | [0.71; 0.87] | 2.0% |
| Dubey et al., 2018 | 125 | 542 | 0.23 | [0.20; 0.27] | 2.0% |
| Mathiyalagen et al., 2019 | 122 | 473 | 0.26 | [0.22; 0.30] | 2.0% |
| Rohatgi et al., 2018 | 9 | 43 | 0.21 | [0.10; 0.36] | 1.9% |
| L. Ahmad et al., 2020 | 30 | 213 | 0.14 | [0.10; 0.19] | 2.0% |
| Goyal et al., 2018 | 129 | 1346 | 0.10 | [0.08; 0.11] | 2.0% |
| Joseph et al., 2020 | 21 | 214 | 0.10 | [0.06; 0.15] | 2.0% |
| Lorenzoni et al., 2019 | 29 | 45 | 0.64 | [0.49; 0.78] | 2.0% |
| Jasti et al., 2018 | 40 | 168 | 0.24 | [0.18; 0.31] | 2.0% |
| Selvaraj & Keshavamurthy, 2016 | 21 | 50 | 0.42 | [0.28; 0.57] | 2.0% |
| **Random effects model** | | **17706** | **0.37** | **[0.28; 0.48]** | **100.0%** |
| **Prediction interval** | | | | **[0.02; 0.94]** | |

Heterogeneity: $I^2 = 98\%$, $\tau^2 = 2.4914$, $p = 0$

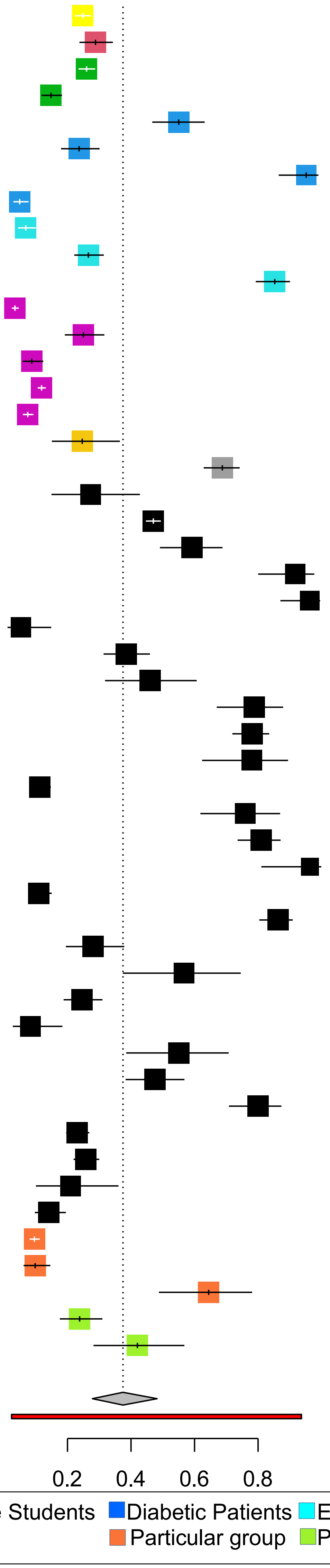
